# Comparing Traditional and Online Problem-Based Learning in Child and Adolescent Psychiatry in Nagoya: A Novel Statistical Approach in Japanese Educational Settings

**DOI:** 10.1101/2025.05.08.25327243

**Authors:** Basile Chretien, Kazuki Nishida, Takeshi Kondo, Noriyuki Takahashi, Hideki Takami, Hiroshi Nishigori, Branko Aleksic, Charles Dolladille, Itzel Bustos Villalobos, Tetsuya Yagi, Norbert Skokauskas, Hideki Kasuya

## Abstract

**Introduction:** Given Japan’s cultural emphasis on group harmony, hierarchical relationships, and indirect communication, the study explores to what degree online Problem-Based Learning (PBL), which requires active participation and self-directed learning, is suitable for Japanese medical students. This is particularly important in fields such as child and adolescent psychiatry, where effective communication and collaborative problem-solving are essential.

**Method:** This study analyses data from medical students at Nagoya University who participated in a Child and Adolescent Psychiatry (CPA) PBL program. In 2019, the program was conducted in-person, while in 2021, another group of students participated in the program online. Both groups completed a 15-item questionnaire. Statistical analyses (regression, factor analysis, clustering) were performed to assess factors influencing satisfaction and engagement with PBL.

**Result:** In-person PBL group reported lower satisfaction (p = 0.002), and younger students and females showed higher satisfaction (p=0.025 and p=0.053, respectively) with PBL. Factor analysis showed two dimensions: Perceived Learning Efficacy and CPA-Specific Appropriateness, with significant group differences (p < 0.001). Cluster analysis identified two groups: Cluster 1 (69% in-person, dissatisfied) and Cluster 2 (57% online, satisfied), with key factors being understanding and enjoyment.

**Discussion:** Online PBL improved student satisfaction, particularly for topics requiring engagement and reflection. These findings suggest that online PBL offers flexibility and accessibility, but challenges related to English language and communication skills remain. Tailoring PBL to specific cultural and demographic contexts, along with fostering international collaboration, could enhance learning outcomes and engagement.

## Introduction

Currently Japan is navigating profound demographic challenges that are reshaping the social and healthcare landscape. With one of the world’s most rapidly aging populations and a consistently declining birth rate, the country is experiencing a demographic inversion that affects every facet of society[1]. The younger generation is shrinking, creating fewer caregivers for the elderly, while the “sandwich generation” of adults, often aged 35–54, is burdened with dual responsibilities: supporting their aging parents and raising their children[2]. These circumstances create a ripple effect on family dynamics and place significant pressure on children and adolescents, who are often caught in the crossfire of these societal and familial stressors[3].

At the same time, modern stressors such as academic pressures, cyberbullying, and the pervasive influence of social media are shaping the mental health landscape of young people[4–6]. Although the absolute number of children in Japan is declining[7], mental health concerns among this population are becoming more prominent[8]. Addressing these challenges requires a robust healthcare system staffed with professionals who are not only clinically skilled but also attuned to the unique sociocultural factors influencing mental health in Japan. Medical students must therefore be equipped to handle such challenges, even if their primary specialization will be outside psychiatry.

A significant barrier to mental healthcare in Japan has historically been the stigma surrounding mental illness[9]. However, societal attitudes are gradually shifting, driven by public health campaigns, improved awareness, and a growing recognition of mental health as a critical component of overall well-being[10]. These changes underscore the importance of preparing future physicians to address mental health issues with competence and empathy. For medical students, early and effective education in child and adolescent psychiatry (CAP) is essential, not only to serve this population but also to support families grappling with complex mental health issues.

In Japan, innovative teaching methods are needed to bridge the gap between theoretical knowledge and practical applications. Problem-Based Learning (PBL), a student-centered pedagogical approach first pioneered at McMaster University in the 1960s, has been heralded as a transformative model in medical education[11]. Unlike traditional lecture-based methods, PBL engages students in active learning through the exploration of real-world clinical scenarios, fostering critical thinking, problem-solving, and collaboration[12]. Over time, PBL has been adopted by medical schools worldwide and has been shown to enhance students’ ability to integrate knowledge and apply it in clinical contexts[13].

In Japan, PBL was introduced relatively late, with the first implementation occurring in 1990 at Tokyo Women’s Medical University. Nagoya University (NU), one of Japan’s premier medical institutions[14] as part of its broader efforts to modernize and globalize medical education. In October 2004, a survey indicated that PBL was the prevalent educational method at 63 of the 79 (80%) Japanese medical schools[15]. Currently, PBL at NU is integrated into the fourth-year medical curriculum, with specific applications in CAP and related disciplines[16]. NU has also partnered with international institutions such as the Norwegian University of Science and Technology (NTNU) to refine its PBL cases and incorporate cross-cultural perspectives, particularly through initiatives like the TroNa Partnership and Equal Partnership.

Despite the growing body of literature on the implementation of PBL, many studies have primarily focused on descriptive outcomes, such as students’ satisfaction or general perceptions of the method using simple correlation tests. While valuable, such analyses fall short of leveraging the full potential of the data generated through PBL programs. Understanding how the underlying pattern of the data in order to try to understand who benefits from the PBL and who does not is very important in order to improve the method locally. Advanced statistical methods like adjusted logistic regression and factorial analyses can uncover patterns and relationships that are not immediately apparent, providing deeper insights into the effectiveness of PBL in fostering essential skills for medical practice[17,18].

This study builds on previously published data on PBL in CAP at NU[19], applying advanced statistical techniques to gain deeper insights and expand upon earlier findings. Adjusted logistic regression and factorial analyses enable a more nuanced exploration of the data. By moving beyond descriptive statistics, this study aims to generate actionable insights that can inform the design and implementation of future PBL programs, both in Japan and globally. The findings have the potential to contribute not only to the academic discourse on PBL but also to the broader field of medical education, where evidence-based approaches are essential in curriculum development.

As Japan continues to adapt to its evolving demographic realities, the importance of training medical professionals who can address the mental health needs of children and adolescents cannot be overstated. The goal of this post-hoc analysis is to compare the traditional, in-person PBL method conducted in English with an online PBL method, also in English, among Japanese medical students. This analysis seeks to determine whether online PBL is suitable for this population, considering their generally limited proficiency in English[20,21], and the cultural tendency toward shyness, particularly when speaking foreign languages [22]. Through this study, we aim to enhance the understanding of PBL’s role in this endeavour, ultimately contributing to the preparation of a healthcare workforce that is equipped to meet the challenges of the future.

## Methods

This study builds upon the data collected from a Problem-Based Learning (PBL) program implemented at Nagoya University (NU) in 2019, focusing on CAP. The dataset consists of student responses to a questionnaire designed to assess their understanding of the course, attitudes toward PBL, and satisfaction with the educational experience. One group conducted PBL in a traditional, in-person setting, while another group participated in PBL in an online format in 2021. The shift to online PBL was initially implemented in response to the COVID-19 pandemic, which necessitated adaptations for remote learning. As this transition occurred naturally and was not randomly assigned, the study follows a naturalistic design rather than a randomized controlled trial. The analysis incorporates a suite of advanced statistical methods to gain deeper insights into the factors that contribute to student success and engagement with the PBL method.

### Data Collection

The original dataset includes responses from fourth-year medical students at NU, who participated in a PBL tutorial on depression in children and adolescents. The tutorial, based on a case involving a hypothetical 15-year-old girl (see Supplementary data 1), was conducted over two 90-minute sessions. Students participated in small groups of 10, each with a designated group leader (Student A) and a note-taker (Student B). The case scenario was presented in English, but students could choose to conduct discussions in either English or Japanese. Following the sessions, students completed a 15-item questionnaire (see Supplementary table 1), which used a three-point Likert scale (1 = agree, 2 = neutral, 3 = disagree) to assess their impressions of the PBL method, specifically in relation to CAP and the case scenario. As questions were all positive, a score of 1 for a question tended to indicate a good appreciation of the PBL, while a score of 3 tended to indicated a bad appreciation of the PBL for this specific point. Missing data were expected to be handled using full case analysis, where only cases with complete data for the variables of interest would be included in the analyses. Descriptive analyses were planned to include all available cases.

## Outcomes

The primary outcome of this study was to evaluate and compare the level of student satisfaction with the PBL educational experience between the traditional in-person delivery and online delivery methods, both conducted in English, after adjusting for potential confounders.

Secondary outcomes included:

1. Identify how questions correlated between each other’s, and what differences could be observed on the correlation between the questions between the 2 groups.
2. Identify the underlying dimensions among the set of questions in order to check that the categorization was appropriately done.
3. Investigate if the result differed between the groups depending on the questions included in the identified dimensions
4. Do a cluster analysis in order to understand how the population can be divided in subgroups depending on their pattern of answers to the survey

### Statistical Analysis

To better understand the data, a series of advanced statistical techniques were employed.

1. Composite Score Development: A composite score was developed to quantify multidimensional aspects of student outcomes. This score was derived from all items in the questionnaire, allowing for a more holistic measure of student satisfaction. The minimum theoretical score was 15 and the maximum theoretical score was 45.
2. A linear regression adjusted on sex and age was done to assess if the composite score depended on the PBL group.
3. Ordered Logistic Regression: Ordered logistic regression was used to analyze the survey responses. This technique allowed us to adjust on age and sex on the likelihood of selecting each Likert scale response (agree, neutral, or disagree) for each question. The ordered nature of the dependent variable was accounted for in this analysis.
4. Visualization of Relationships: Pairwise scatter plots and correlation heatmaps were generated to explore relationships between the variables. These visualizations provide an intuitive understanding of the interactions between variables, including how different aspects of the students’ experiences were related.
5. Factorial Analysis: Exploratory Factor Analysis (EFA) was conducted to identify latent dimensions underlying the survey responses. Principal axis factoring with varimax rotation was applied, retaining factors with eigenvalues greater than 1. This analysis aimed to determine whether distinct dimensions of student engagement emerged, such as communication skills development, content understanding, and self-directed learning.
6. Clustering Analysis: To identify natural groupings of students based on their responses, k-means clustering was performed. The number of clusters was determined using the elbow method (WSS) and the silhouette method. After selecting the optimal number of clusters, k-means clustering was applied, and each student was assigned a cluster label. Cluster profiles were summarized by calculating the mean and standard deviation of responses for each question. Differences between clusters were tested using p-values. A random forest analysis was also performed to assess the importance of specific survey responses in predicting cluster membership.

R 4.4.1 software was used for all statistical analyses[23]. Descriptive statistics were first calculated for the sample, followed by the application of advanced statistical techniques such as ordered logistic regression, factorial analysis, and clustering analysis. Pearson’s pairwise correlations were calculated to assess the relationships between variables, with statistical significance set at p ≤ 0.05.

### Ethical Considerations

All procedures were conducted in accordance with the ethical standards of Nagoya University and the 1964 Helsinki Declaration and its later amendments. The data used in this study were originally collected as part of a routine educational evaluation of a PBL program. Participation in the questionnaire was voluntary. Students were informed that their anonymous responses might be used for quality improvement and future academic purposes, and that they could oppose the use of their data without any consequences. Verbal consent was obtained from all participants prior to data collection. This consent process was carried out by the instructors during the session, and agreement to participate was documented through the submission of the completed questionnaire. Formal ethical approval for the retrospective analysis of the anonymized dataset for research purposes was obtained from the Nagoya University Ethical Committee on March 5th, 2022 (Approval Code: 2021-0482). The dataset was accessed for research purposes on March 22^nd^, 2022.

## Results

Of the 112 students who participated in the in-person PBL, 109 completed the survey, while among the 111 participants in online setting, 100 participants filled out the questionnaire. Age ranged from 21 to 32 years old and was not statistically different among the 2 groups with a median of 22 years old for both groups. Males were more represented than females in the 2 groups (76% in the classic group and 73% in the online group) which was not statistically different. Except one Chinese student in the online group, all other students were Japanese (Table 1).

**Table 1.** Demographics.

| **PBL type** | **N** | **classic N = 109<sup>1</sup>** | **online N = 100<sup>1</sup>** | **p-value<sup>2</sup>** |
| --- | --- | --- | --- | --- |
| **Age** | 191 | 22.00 (22.00, 23.00) | 22.00 (22.00, 23.00) | 0.6 |
| Unknown |  | 11 | 7 |  |
| **Gender** | 189 |  |  | 0.6 |
| F |  | 23 (24%) | 25 (27%) |  |
| M |  | 74 (76%) | 67 (73%) |  |
| Unknown |  | 12 | 8 |  |
| **Nationality** | 202 |  |  | 0.5 |
| 1 |  | 109 (100%) | 92 (99%) |  |
| 2 (Chinese) |  | 0 (0%) | 1 (1.1%) |  |
| Unknown |  | 0 | 7 |  |

<sup>1</sup>Median (Q1, Q3); n (%)
<sup>2</sup>Wilcoxon rank sum test; Pearson's Chi-squared test; Fisher's exact test

Questionnaire results are shown in Table 2, and students in the online group gave a significantly better appreciation of the PBL compared to the in-person group (Table 2). Similar results are found with the ordered logistic regression after adjusting on age and sex (Table 3). Results of the composite score are presented in Figure 1A and were statistically higher in the in-person group according to the multiple linear regression (p = 0.02) (Table 4). Since the original data did not follow a normal distribution (skewness = 0.77, kurtosis = 2.71), we acknowledge a potential violation of the normality assumption. However, the residuals from the linear regression were approximately normal (skewness = 0.77, kurtosis = 3.02), supporting the validity of the analysis under the assumption of normal residuals (Supplementary Figure 1). Despite the non-normality of the original data, multiple linear regression remains a robust method in this context[24]. Age was significantly associated with a higher composite score, indicating lower satisfaction among older students. A trend toward higher scores in males was also observed, though not statistically significant. Additionally, a higher proportion of students in the online group reported very high appreciation of PBL, whereas the in-person group had a lower proportion of highly satisfied students.

**Figure 1:**
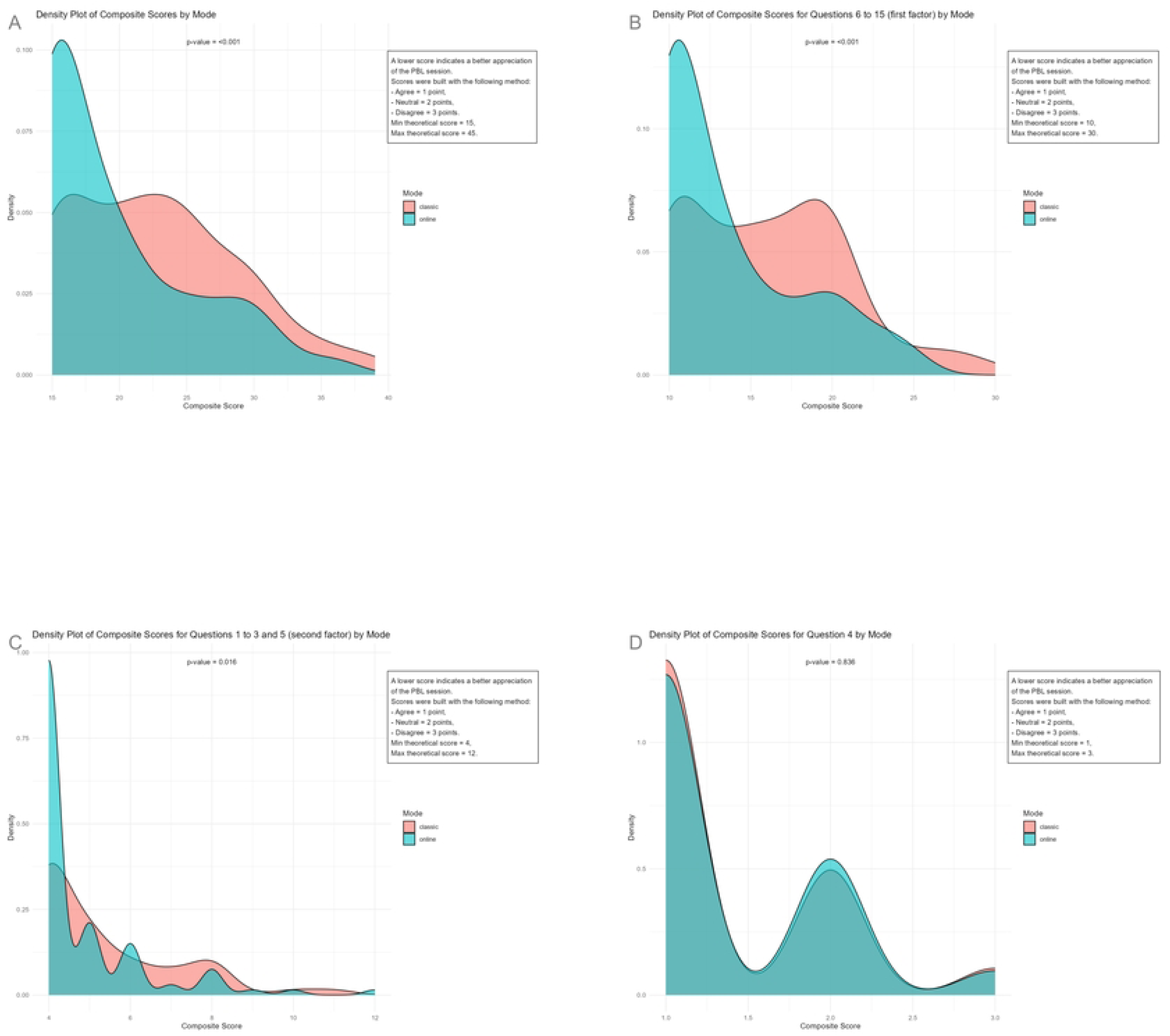
Density plot of Composite score by mode (A: for the whole questionnaire, B: for the 1^st^ dimension by Mode, C: for the 2^nd^ dimension by Mode, D: for the 4^th^ question)

**Table 2.** Answers to the satisfaction questionnaire after PBL.

| **Table 2. Answers to the satisfaction questionnaire after PBL** |  |  |  |  |
| --- | --- | --- | --- | --- |
| **PBL type** | **N** | **classic N**<br>= 109<sup>1</sup> | **online N**<br>= 100<sup>1</sup> | **p-value<sup>2</sup>** |
| **Q1 : PBL helps me to develop skills to solve clinical problems** | 208 |  |  | 0.078 |
| 1 |  | 89 (82%) | 91 (92%) |  |
| 2 |  | 17 (16%) | 6 (6.1%) |  |
| 3 |  | 3 (2.8%) | 2 (2.0%) |  |
| Unknown |  | 0 | 1 |  |
| **Q2 : PBL is a good way of learning the content of the course** | 208 |  |  | 0.034 |
| 1 |  | 77 (71%) | 85 (86%) |  |
| 2 |  | 27 (25%) | 12 (12%) |  |
| 3 |  | 5 (4.6%) | 2 (2.0%) |  |
| Unknown |  | 0 | 1 |  |
| **Q3 : PBL encourages my self-directed learning** | 208 |  |  | 0.040 |
| 1 |  | 71 (65%) | 80 (81%) |  |
| 2 |  | 33 (30%) | 17 (17%) |  |
| 3 |  | 5 (4.6%) | 2 (2.0%) |  |
| Unknown |  | 0 | 1 |  |
| **Q4 : PBL helps to develop my communication skills** | 208 |  |  | >0.900 |
| 1 |  | 75 (69%) | 66 (67%) |  |
| 2 |  | 28 (26%) | 28 (28%) |  |
| 3 |  | 6 (5.5%) | 5 (5.1%) |  |
| Unknown |  | 0 | 1 |  |
| **Q5 : I think, PBL is more effective way of learning compared to traditional lecture** | 208 |  |  | 0.600 |
| 1 |  | 71 (65%) | 71 (72%) |  |
| 2 |  | 33 (30%) | 24 (24%) |  |
| 3 |  | 5 (4.6%) | 4 (4.0%) |  |
| Unknown |  | 0 | 1 |  |
| **Q6 : PBL enhanced my understanding about child & adolescent psychiatry** | 208 |  |  | 0.001 |
| 1 |  | 58 (53%) | 76 (77%) |  |
| 2 |  | 40 (37%) | 20 (20%) |  |
| 3 |  | 11 (10%) | 3 (3.0%) |  |
| Unknown |  | 0 | 1 |  |
| **Q7 : PBL increased my interest in child & adolescent psychiatry** | 208 |  |  | 0.300 |
| 1 |  | 60 (55%) | 65 (66%) |  |
| 2 |  | 36 (33%) | 26 (26%) |  |
| 3 |  | 13 (12%) | 8 (8.1%) |  |
| Unknown |  | 0 | 1 |  |
| **Q8 : Child and adolescent psychiatry because of it's emphasis on teamwork and collaboration, would be a specialty learned optimally through PBL** | 208 |  |  | 0.092 |
| 1 |  | 48 (44%) | 56 (57%) |  |
| 2 |  | 50 (46%) | 39 (39%) |  |
| 3 |  | 11 (10%) | 4 (4.0%) |  |
| Unknown |  | 0 | 1 |  |
| **Q9 : Duration of PBL session was enough to learn about child & adolescent psychiatry** | 208 |  |  | 0.300 |
| 1 |  | 50 (46%) | 56 (57%) |  |
| 2 |  | 41 (38%) | 32 (32%) |  |
| 3 |  | 18 (17%) | 11 (11%) |  |
| Unknown |  | 0 | 1 |  |
| **Q10 : I enjoyed learning about child and adolescent psychiatry via PBL** | 205 |  |  | <0.001 |
| 1 |  | 48 (44%) | 68 (71%) |  |
| 2 |  | 47 (43%) | 22 (23%) |  |
| 3 |  | 14 (13%) | 6 (6.3%) |  |
| Unknown |  | 0 | 4 |  |
| **Q11 : I think Mayumi's case is common for a clinical-settings in Japan.** | 208 |  |  | 0.500 |
| 1 |  | 61 (56%) | 57 (58%) |  |
| 2 |  | 40 (37%) | 31 (31%) |  |
| 3 |  | 8 (7.3%) | 11 (11%) |  |
| Unknown |  | 0 | 1 |  |
| **Q12 : Mayumi's case was well written and understandable** | 208 |  |  | 0.004 |
| 1 |  | 60 (55%) | 76 (77%) |  |
| 2 |  | 40 (37%) | 20 (20%) |  |
| 3 |  | 9 (8.3%) | 3 (3.0%) |  |
| Unknown |  | 0 | 1 |  |
| **Q13 : Mayumi's case has an interesting clinical trigger** | 208 |  |  | 0.002 |
| 1 |  | 44 (40%) | 64 (65%) |  |
| 2 |  | 53 (49%) | 29 (29%) |  |
| 3 |  | 12 (11%) | 6 (6.1%) |  |
| Unknown |  | 0 | 1 |  |
| **Q14 : Mayumi's case has an appropriate level of difficulty/challenge** | 208 |  |  | 0.002 |
| 1 |  | 58 (53%) | 76 (77%) |  |
| 2 |  | 43 (39%) | 20 (20%) |  |
| 3 |  | 8 (7.3%) | 3 (3.0%) |  |
| Unknown |  | 0 | 1 |  |
| **Q15 : Mayumi's case helped me to understand the importance of multidisciplinary approach in child and adolescent psychiatry** | 207 |  |  | 0.009 |
| 1 |  | 62 (57%) | 75 (76%) |  |
| 2 |  | 39 (36%) | 23 (23%) |  |
| 3 |  | 7 (6.5%) | 1 (1.0%) |  |
| Unknown |  | 1 | 1 |  |

<sup>1</sup>Median (Q1, Q3); n (%)
<sup>2</sup>Wilcoxon rank sum test; Pearson's Chi-squared test; Fisher's exact test

**Table 3.** ordered logistic regression adjusted on age and sex in order to compare the satisfaction responses between groups to the questionnaire about PBL.

|  |  |  |  |  |  |  |  |  |  |  |  |  |  |  |  |
| --- | --- | --- | --- | --- | --- | --- | --- | --- | --- | --- | --- | --- | --- | --- | --- |
| Predictors | factor(Q1) |  |  | factor(Q2) |  |  | factor(Q3) |  |  | factor(Q4) |  |  | factor(Q5) |  |  |
|  | Odds Ratios | CI | p | Odds Ratios | CI | p | Odds Ratios | CI | p | Odds Ratios | CI | p | Odds Ratios | CI | p |
| classic | Referenc<br>e |  |  | Referenc<br>e |  |  | Referenc<br>e |  |  | Referenc<br>e |  |  | Referenc<br>e |  |  |
| online | 0.46 | 0.18 – 1.10 | 0.094 | 0.43 | 0.20 – 0.89 | **0.027** | 0.51 | 0.26 – 0.98 | **0.047** | 1.18 | 0.65 – 2.17 | 0.587 | 0.78 | 0.41 – 1.46 | 0.441 |
| Age | 1.04 | 0.76 – 1.33 | 0.778 | 1.21 | 0.99 – 1.47 | 0.055 | 1.02 | 0.81 – 1.25 | 0.831 | 0.93 | 0.73 – 1.13 | 0.483 | 1.39 | 1.15 – 1.70 | **0.001** |
| F | Referenc<br>e |  |  | Referenc<br>e |  |  | Referenc<br>e |  |  | Referenc<br>e |  |  | Referenc<br>e |  |  |
| M | 1.38 | 0.52 – 4.36 | 0.550 | 1.70 | 0.72 – 4.51 | 0.250 | 2.34 | 1.05 – 5.79 | **0.049** | 1.11 | 0.57 – 2.25 | 0.761 | 1.30 | 0.64 – 2.79 | 0.478 |
| Observations | 188 |  |  | 188 |  |  | 188 |  |  | 188 |  |  | 188 |  |  |
| Predictors | factor(Q6) |  |  | factor(Q7) |  |  | factor(Q8) |  |  | factor(Q9) |  |  | factor(Q10) |  |  |
|  | Odds Ratios | CI | p | Odds Ratios | CI | p | Odds Ratios | CI | p | Odds Ratios | CI | p | Odds Ratios | CI | p |
| classic | Referenc<br>e |  |  | Referenc<br>e |  |  | Referenc<br>e |  |  | Referenc<br>e |  |  | Referenc<br>e |  |  |
| online | 0.36 | 0.19 – 0.67 | **0.002** | 0.68 | 0.38 – 1.22 | 0.203 | 0.65 | 0.37 – 1.15 | 0.141 | 0.71 | 0.41 – 1.22 | 0.218 | 0.36 | 0.20 – 0.66 | **0.001** |
| Age | 1.10 | 0.91 – 1.33 | 0.306 | 1.15 | 0.97 – 1.37 | 0.097 | 1.32 | 1.10 – 1.61 | **0.004** | 1.14 | 0.95 – 1.36 | 0.153 | 1.34 | 1.11 – 1.63 | **0.002** |
| F | *Reference*<br>*e* |  |  | *Reference*<br>*e* |  |  | *Reference*<br>*e* |  |  | *Reference*<br>*e* |  |  | *Reference*<br>*e* |  |  |
| M | 1.98 | 0.96 – 4.31 | 0.074 | 2.22 | 1.12 – 4.67 | **0.029** | 1.72 | 0.89 – 3.38 | 0.113 | 1.32 | 0.71 – 2.51 | 0.383 | 2.35 | 1.17 – 4.94 | **0.021** |
| Observations | 188 |  |  | 188 |  |  | 188 |  |  | 188 |  |  | 185 |  |  |

|  | factor(Q11) |  |  | factor(Q12) |  |  | factor(Q13) |  |  | factor(Q14) |  |  | factor(Q15) |  |  |
| --- | --- | --- | --- | --- | --- | --- | --- | --- | --- | --- | --- | --- | --- | --- | --- |
| *Predictors* | *Odds Ratios* | *CI* | *p* | *Odds Ratios* | *CI* | *p* | *Odds Ratios* | *CI* | *p* | *Odds Ratios* | *CI* | *p* | *Odds Ratios* | *CI* | *p* |
| classic | *Reference*<br>*e* |  |  | *Reference*<br>*e* |  |  | *Reference*<br>*e* |  |  | *Reference*<br>*e* |  |  | *Reference*<br>*e* |  |  |
| online | 1.07 | 0.60 – 1.90 | 0.824 | 0.37 | 0.19 – 0.69 | **0.003** | 0.45 | 0.25 – 0.80 | **0.008** | 0.37 | 0.20 – 0.69 | **0.002** | 0.51 | 0.27 – 0.94 | **0.035** |
| Age | 1.17 | 0.98 – 1.40 | 0.083 | 1.18 | 0.97 – 1.43 | 0.097 | 1.17 | 0.97 – 1.41 | 0.099 | 1.08 | 0.89 – 1.31 | 0.416 | 1.04 | 0.83 – 1.29 | 0.700 |
| F | *Reference*<br>*e* |  |  | *Reference*<br>*e* |  |  | *Reference*<br>*e* |  |  | *Reference*<br>*e* |  |  | *Reference*<br>*e* |  |  |
| M | 2.47 | 1.24 – 5.20 | **0.014** | 1.16 | 0.57 – 2.45 | 0.688 | 0.84 | 0.44 – 1.59 | 0.580 | 1.70 | 0.82 – 3.71 | 0.170 | 1.85 | 0.89 – 4.11 | 0.116 |
| Observations | 188 |  |  | 188 |  |  | 188 |  |  | 188 |  |  | 187 |  |  |

**Table 4:** Result of the multiple linear regression to compare the composite score between the two groups.

| Variable | Estimate | 95%CI | p-value |
| --- | --- | --- | --- |
| Classic (reference) | - | - | - |
| Online | -2.66 | [-4.36 ; -0.97] | 0.002 |
| Age | 0.64 | 0.08-1.20 | 0.025 |
| F (reference) | - | - | - |
| M | 1.90 | [-0.03 ; 3.83] | 0.053 |

When exploring the relationship between different questions, the pairwise scatter plots revealed a consistent response pattern among students. Specifically, students who selected a high rating (e.g., 1) on the Likert scale for one question tended to give similarly high ratings to other related questions. This pattern suggests a strong correlation between certain responses (Supplementary Figures 2, 3, and 4).

The correlation matrix found high correlations between question 1 to 3, 6 to 10 and 12 to 15. Lower correlations were found between question 4 and the other questions, as well as between question 11 and the other questions. A negative correlation between question 4 and question 12 was found only in the classic group (Supplementary Figures 5, 6 and 7). A scree plot was done in order to assess the number of dimensions for the factor analysis (Figure 1B). A factor analysis was done for 2 dimensions (Table 5), and questions 6 to 15 were associated to the first dimension, while questions 1, 2, 3 and 5 were associated to the second dimension. Question 4 was not associated with any dimension (Figure 2A). The composite score based on the factor analysis for the first dimension was also significantly different between the 2 groups. The density plot also found a high satisfaction of the students on the online group compared to the classic group (Figure 2B). A similar result was observed for the second dimension (Figure 2C), while almost no difference could be observed between the groups for the question 4 (Figure 2D).

**Figure 2:**
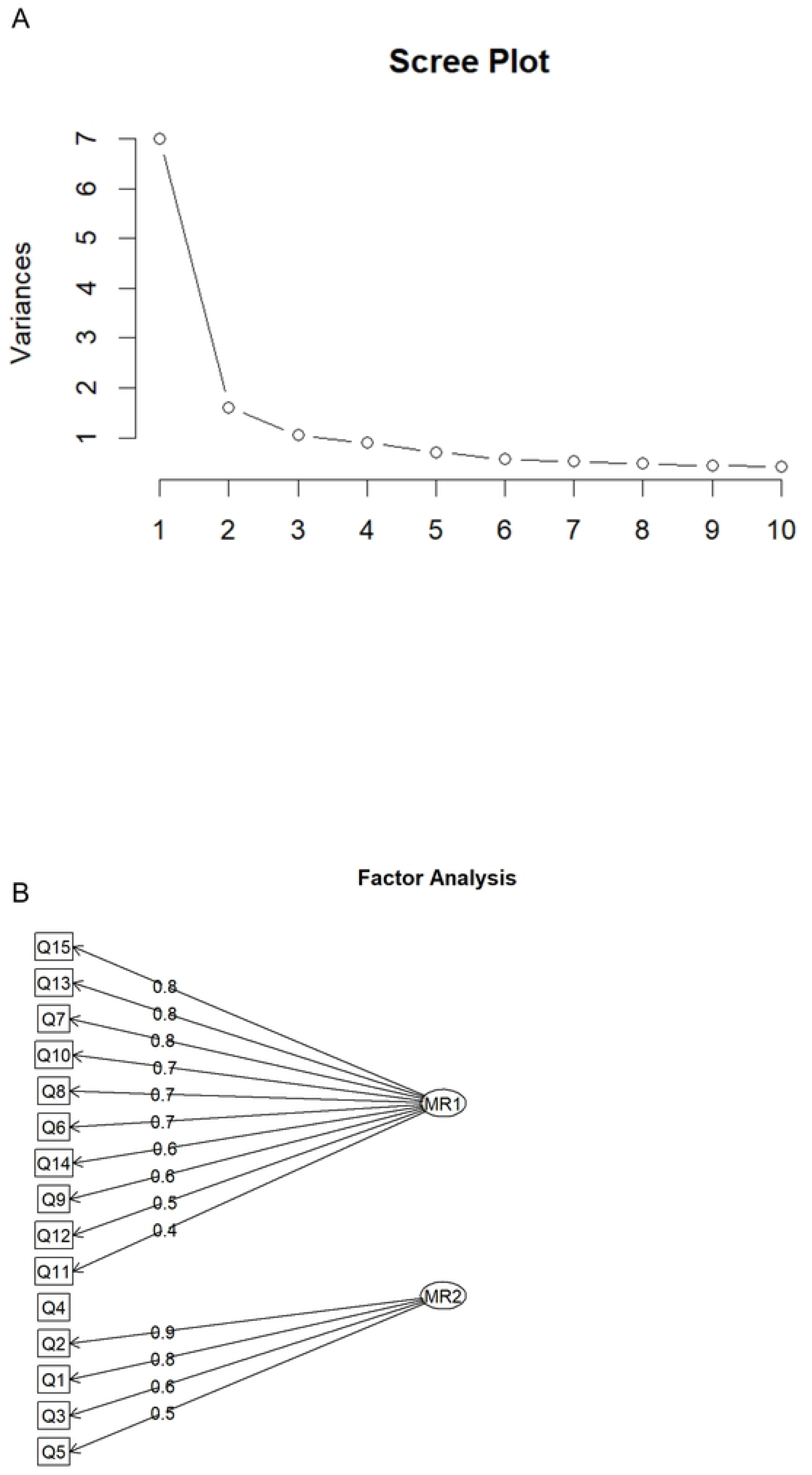
Factor analysis (A: Scree plot, B: diagram)

**Table 5.** Results of the Factor Analysis.

| **Variable** | **Factor 1 Loadings** | **Factor 2 Loadings** | **Uniqueness** |
| --- | --- | --- | --- |
| Q1 | 0.1449725 | 0.7782357 | 0.3733321 |
| Q2 | 0.1722254 | 0.8676928 | 0.2174476 |
| Q3 | 0.3021317 | 0.6236074 | 0.5198303 |
| Q4 | 0.2992293 | 0.2952419 | 0.8232940 |
| Q5 | 0.3675036 | 0.5017134 | 0.6132247 |
| Q6 | 0.7291871 | 0.3714524 | 0.3303093 |
| Q7 | 0.7554548 | 0.2605440 | 0.3614049 |
| Q8 | 0.7321041 | 0.2964379 | 0.3761482 |
| Q9 | 0.6294869 | 0.3653332 | 0.4702779 |
| Q10 | 0.7414557 | 0.3032310 | 0.3582943 |
| Q11 | 0.4438675 | 0.2187698 | 0.7551214 |
| Q12 | 0.5298896 | 0.1904392 | 0.6829499 |
| Q13 | 0.7573570 | 0.2227423 | 0.3767963 |
| Q14 | 0.6467732 | 0.0850467 | 0.5744515 |
| Q15 | 0.7577603 | 0.1735414 | 0.3956827 |

The key characteristics of each cluster are presented in Table 6. The cluster plot indicated a good separation of the students (Supplementary Figure 8). Cluster 1 is composed of 77 students, and is mostly composed of students belonging to the in-person group (69%), and is also composed of 85% of male students and present high proportions of “neutral” or “disagree” answers, indicating a dissatisfaction of the students of this group toward PBL. By contrast, Cluster 2 is composed of 127 students, with a majority of students belonging to the online group (57%). The proportion of males is lower in this cluster (67%) and it presents a very high proportion of “agree” answers, indicating a strong satisfaction of the students in this group toward PBL.

**Table 6.** Results of the clustering analysis.

|  |  | **1** | **2** | **p-** |
| --- | --- | --- | --- | --- |
| **By Cluster** | **N** | N = 77<sup>1</sup> | N = 127<sup>1</sup> | **value**<sup>2</sup> |
| **Mode** | 204 |  |  | <0.001 |
| classic |  | 53 (69%) | 55 (43%) |  |
| online |  | 24 (31%) | 72 (57%) |  |
| **Age** | 186 | 22.00<br>(22.00,<br>23.00) | 22.00<br>(21.50,<br>23.00) | 0.006 |
| Unknown |  | 7 | 11 |  |
| **Gender** | 184 |  |  | 0.007 |
| F |  | 10 (15%) | 38 (33%) |  |
| M |  | 58 (85%) | 78 (67%) |  |
| Unknown |  | 9 | 11 |  |
| **Nationality** | 197 |  |  | >0.900 |
| 1 |  | 76 (100%) | 120 (99%) |  |
| 2 (Chinese) |  | 0 (0%) | 1 (0.8%) |  |
| Unknown |  | 1 | 6 |  |
| **Q1 : PBL helps me to develop skills to solve clinical problems** | 204 |  |  | <0.001 |
| 1 |  | 54 (70%) | 123 (97%) |  |
| 2 |  | 18 (23%) | 4 (3.1%) |  |
| 3 |  | 5 (6.5%) | 0 (0%) |  |

**Table 6. Results of the clustering analysis**
|  |  | **1** | **2** | **p-** |
| --- | --- | --- | --- | --- |
| **By Cluster** | **N** | **N = 77<sup>1</sup>** | **N = 127<sup>1</sup>** | **value<sup>2</sup>** |
| **Q2 : PBL is a good way of learning the content of the course** | 204 |  |  | <0.001 |
| 1 |  | 41 (53%) | 118 (93%) |  |
| 2 |  | 29 (38%) | 9 (7.1%) |  |
| 3 |  | 7 (9.1%) | 0 (0%) |  |
| **Q3 : PBL encourages my self-directed learning** | 204 |  |  | <0.001 |
| 1 |  | 35 (45%) | 115 (91%) |  |
| 2 |  | 35 (45%) | 12 (9.4%) |  |
| 3 |  | 7 (9.1%) | 0 (0%) |  |
| **Q4 : PBL helps to develop my communication skills** | 204 |  |  | <0.001 |
| 1 |  | 38 (49%) | 102 (80%) |  |
| 2 |  | 30 (39%) | 23 (18%) |  |
| 3 |  | 9 (12%) | 2 (1.6%) |  |
| **Q5 : I think, PBL is more effective way of learning compared to traditional lecture** | 204 |  |  | <0.001 |
| 1 |  | 28 (36%) | 113 (89%) |  |
| 2 |  | 42 (55%) | 12 (9.4%) |  |
| 3 |  | 7 (9.1%) | 2 (1.6%) |  |
| **Q6 : PBL enhanced my understanding about child & adolescent psychiatry** | 204 |  |  | <0.001 |

**Table 6. Results of the clustering analysis**
|  |  | **1** | **2** | **p-** |
| --- | --- | --- | --- | --- |
| **By Cluster** | **N** | **N = 77<sup>1</sup>** | **N = 127<sup>1</sup>** | **value<sup>2</sup>** |
| 1 |  | 11 (14%) | 119 (94%) |  |
| 2 |  | 52 (68%) | 8 (6.3%) |  |
| 3 |  | 14 (18%) | 0 (0%) |  |
| **Q7 : PBL increased my interest in child & adolescent psychiatry** | 204 |  |  | <0.001 |
| 1 |  | 13 (17%) | 111 (87%) |  |
| 2 |  | 44 (57%) | 15 (12%) |  |
| 3 |  | 20 (26%) | 1 (0.8%) |  |
| **Q8 : Child and adolescent psychiatry because of it's emphasis on teamwork and collaboration, would be a specialty learned optimally through PBL** | 204 |  |  | <0.001 |
| 1 |  | 4 (5.2%) | 99 (78%) |  |
| 2 |  | 60 (78%) | 26 (20%) |  |
| 3 |  | 13 (17%) | 2 (1.6%) |  |
| **Q9 : Duration of PBL session was enough to learn about child & adolescent psychiatry** | 204 |  |  | <0.001 |
| 1 |  | 7 (9.1%) | 98 (77%) |  |
| 2 |  | 44 (57%) | 26 (20%) |  |
| 3 |  | 26 (34%) | 3 (2.4%) |  |
| **Q10 : I enjoyed learning about child and adolescent psychiatry via PBL** | 204 |  |  | <0.001 |

**Table 6. Results of the clustering analysis**
|  |  | **1** | **2** | **p-** |
| --- | --- | --- | --- | --- |
| **By Cluster** | **N** | **N = 77<sup>1</sup>** | **N = 127<sup>1</sup>** | **value<sup>2</sup>** |
| 1 |  | 7 (9.1%) | 108 (85%) |  |
| 2 |  | 50 (65%) | 19 (15%) |  |
| 3 |  | 20 (26%) | 0 (0%) |  |
| **Q11 : I think Mayumi's case is common for a clinical-settings in Japan.** | 204 |  |  | <0.001 |
| 1 |  | 19 (25%) | 98 (77%) |  |
| 2 |  | 45 (58%) | 26 (20%) |  |
| 3 |  | 13 (17%) | 3 (2.4%) |  |
| **Q12 : Mayumi's case was well written and understandable** | 204 |  |  | <0.001 |
| 1 |  | 23 (30%) | 112 (88%) |  |
| 2 |  | 44 (57%) | 13 (10%) |  |
| 3 |  | 10 (13%) | 2 (1.6%) |  |
| **Q13 : Mayumi's case has an interesting clinical trigger** | 204 |  |  | <0.001 |
| 1 |  | 8 (10%) | 99 (78%) |  |
| 2 |  | 55 (71%) | 27 (21%) |  |
| 3 |  | 14 (18%) | 1 (0.8%) |  |
| **Q14 : Mayumi's case has an appropriate level of difficulty/challenge** | 204 |  |  | <0.001 |
| 1 |  | 27 (35%) | 106 (83%) |  |

**Table 6. Results of the clustering analysis**
| By Cluster | N | 1 | 2 | p-value<sup>2</sup> |
| --- | --- | --- | --- | --- |
|  |  | N = 77<sup>1</sup> | N = 127<sup>1</sup> |  |
| 2 |  | 40 (52%) | 20 (16%) |  |
| 3 |  | 10 (13%) | 1 (0.8%) |  |
| **Q15 : Mayumi's case helped me to understand the importance of multidisciplinary approach in child and adolescent psychiatry** | 204 |  |  | <0.001 |
| 1 |  | 20 (26%) | 117 (92%) |  |
| 2 |  | 50 (65%) | 9 (7.1%) |  |
| 3 |  | 7 (9.1%) | 1 (0.8%) |  |
| <sup>1</sup> n (%); Median (Q1, Q3) |  |  |  |  |
| <sup>2</sup> Pearson's Chi-squared test; Wilcoxon rank sum test; Fisher's exact test |  |  |  |  |

The feature importance for cluster identification indicated that question 6 (PBL enhanced my understanding of child and adolescent psychiatry) and question 10 (I enjoyed learning about child and adolescent psychiatry via PBL) were the most important (Supplementary Figure 9).

## Discussion

Our study aimed to compare the appreciation and outcomes of in-person and online PBL sessions in CAP. The findings indicate significant differences in the students’ perceptions and satisfaction levels between the two modalities, with implications for the future design and delivery of PBL.

### Principal Findings

The primary observation was a higher overall satisfaction reported by students in the online PBL group compared to the in-person group. This was evidenced by the direct questionnaire responses (Table 2), the ordered logistic regression analysis, adjusted for age and sex (Table 3), and the results of the cluster analyses (Table 6). The composite scores derived from factor analysis further highlighted nuanced differences, with higher satisfaction levels in the online group for dimensions associated with questions 6 to 15, which focused on the perceived educational value and enjoyment of PBL (Figure 2B).

Interestingly, the in-person group demonstrated significantly higher composite scores derived from multiple linear regression (p = 0.002, Table 4). These higher scores, indicating lower satisfaction, were associated with older age and a trend toward male predominance, suggesting that certain demographic factors may moderate PBL satisfaction. Additionally, the clustering analysis revealed distinct profiles: Cluster 1, dominated by in-person participants, exhibited higher dissatisfaction, whereas Cluster 2, primarily comprising online participants, demonstrated strong satisfaction (Table 6).

### Interpretation of Findings

The high satisfaction reported in the online PBL group aligns with emerging literature emphasizing the advantages of digital learning environments, particularly during the COVID-19 pandemic. Studies have noted that online platforms enable greater flexibility, accessibility, and personalized learning experiences [25,26]. In a qualitative study performed in 2004, students with experience in in-person PBL felt that online PBL increased their flexibility for learning, enhanced their ability to deeply process content, and provided access to valuable learning resources. However, they also experienced a period of adaptation to the online environment, perceived a heavy workload, and had difficulties making group decisions online, probably because of the old technology used for the PBL platform [27]. Our results further suggest that online PBL may enhance engagement and appreciation for topics like child and adolescent psychiatry, as reflected by the importance of questions 6 and 10 in cluster identification.

In contrast, the lower satisfaction in the in-person group may stem from logistical challenges, rigid schedules, or reduced opportunities for individual reflection. Additionally, the fact that the PBL sessions were conducted in English might have contributed to the preference for online PBL, especially given the Japanese students’ potential need for additional time and resources to process and articulate their thoughts in a non-native language. This challenge is not unique to Japan and could similarly affect students from other non-English-speaking countries participating in English-medium PBL sessions. Online environments may have provided a more comfortable and less pressured setting, enabling students to review materials, formulate responses, and participate at their own pace. These findings are consistent with previous research highlighting the importance of learner autonomy and the role of language accessibility in PBL environments [28]. This is one of the reasons why authors of a systematic review about adoption of PBL in medical schools in non-western countries have recently recommended to proactively consider ways to adapt PBL education taking into account local contexts and culture [28]. Additionally, the observed pattern of correlated responses among certain questionnaire items (e.g., questions 1-3, 6-10, and 12-15) underscores the structured nature of PBL, where interconnected learning outcomes may influence students’ overall perceptions.

When building the questionnaire, we divided it into 3 sections: the first section (questions 1 to 5) called “General impressions of PBL”, the second section (questions 6 to 10) called “Questions about PBL and child and adolescent psychiatry” and the third section (questions 11 to 16) called “Specific questions about the case”. The Factor analysis allowed us to understand that the second and third sections were included in a single dimension, which is consistent with their purposes. Four out of five questions included in the first section were included in the second dimension. The only question not linked to any dimension in this questionnaire was the fourth question: “PBL helps develop my communication skills”. Percentages of answers in each category of the Likert scale were extremely close among the two groups (p = 0.59 after adjustment) and were not statistically significantly impacted by neither age nor sex. A high proportion of students agreed with this question in both groups (69% in the in-person group and 67% in the online group). To enhance communication among healthcare professionals—a central objective of PBL [11]—it is essential to consider the challenges faced by students with high levels of communication apprehension. These students may experience significant difficulties participating in PBL sessions [29]. Therefore, when designing PBL scenarios and defining their implementation context, particular attention should be given to creating systems that facilitate communication opportunities for students with such apprehension.

### Implications for Practice

These findings have several implications for teaching about CAP and medical education overall. First, transitioning to or incorporating online PBL can improve student satisfaction, particularly for complex topics requiring active participation and reflection. Second, demographic factors such as age and gender should be considered in PBL design, with targeted interventions to address potential disparities. For example, younger students and females in our study showed relatively higher satisfaction. This observation aligns with broader cultural trends in Japan, where women often exhibit a heightened interest in foreign cultures and languages [30,31].

Such interests might make them more receptive to English-language PBL, especially in an online format that allows greater exposure to and interaction with international educational methods. This trend underscores the need for tailored approaches to enhance engagement among older and male participants, who may not share the same level of intrinsic motivation for language and cultural immersion. One potential solution is to design scenarios in which the patient is a foreigner or the students are participating in internships in English-speaking countries, or Japanese tourists gets sick overseas. This approach enhances the sense of immersion and emphasizes the necessity of English[32]. Moreover, a study involving 48 Catalan/Spanish students who studied abroad demonstrated that they improved their English skills to a similar extent, regardless of whether they studied in an anglophone or non-anglophone country [33]. Thus, we propose that employing PBL in English for non-native English speakers could not only enhance medical education but also promote language acquisition. Collaboration between universities across different countries is likely to further increase student engagement and knowledge retention, as it exposes students to diverse perspectives, fosters cross-cultural communication, and creates opportunities for peer learning in a global context [34].

The slightly negative correlation (−0.09) observed between question 4 (“PBL helps to develop my communication skills”) and question 12 (“Mayumi’s case was well-written and understandable”) in the in-person group may indicate that these students faced greater challenges accessing translation tools, which could have hindered their understanding of the scenario compared to the online group. However, this limitation does not appear to have diminished the effectiveness of PBL in enhancing their communication skills. The lack of association of question 4 (“PBL helped me develop my communication skills”) with any dimension in the factor analysis warrants further investigation. Communication is a core competency in medical education (4), and its low-correlation with assessment of PBL efficacy may be influenced by the specific scenario used in this study. Testing different scenarios could help identify whether scenario design affects the development of communication skills and inform curriculum revisions to better address this essential competency. The most important questions in order to be classed in the cluster analysis were questions 6 (PBL enhanced my understanding about child and adolescent psychiatry) and question 10 (I enjoyed learning about child and adolescent psychiatry via PBL). This suggests a strong preference among participants for engaging, enjoyable learning experiences, highlighting the importance of ludic elements in education. This is consistent with the results of a Brazilian study on 106 veterinary medicine students, which showed that gamed-based learning allows knowledge acquirement in the same scale as case-based learning (CBL) and peer tutoring, but also promotes a greater perception of the stimulus for self-study and problem-solving ability and contributes to the development of group dynamics compared with the group who received CBL (P < 0.05) [35].

### Strengths and Limitations

Our study is among the first to employ both factor analysis and clustering to evaluate PBL satisfaction comprehensively. The inclusion of multiple statistical approaches, including logistic regression, linear regression, factor analyses and cluster analyses, adds robustness to our findings. Additionally, the use of a standardized questionnaire allowed for detailed exploration of student perceptions across different dimensions.

However, several limitations should be acknowledged. First, the study population was predominantly Japanese, limiting the generalizability of our findings to other cultural contexts. Additionally, the study did not assess the English proficiency of participants prior to the PBL sessions. Research indicates that Japanese students often face challenges in expressing themselves and comprehending content in English due to differences in linguistic structures and limited opportunities for conversational practice [20]. These factors may have contributed to the findings. To address these issues, future implementations should consider providing tailored vocabulary lists, glossaries, and other language aids designed to align with the specific PBL topics and collaborate with international universities. Encouraging participation through structured prompts and fostering a supportive, low-pressure environment may also help mitigate language barriers and enhance the inclusivity of PBL sessions. Third, the cross-sectional design precludes causal inference, and longitudinal studies are needed to assess the long-term impact of PBL modalities on educational outcomes. Fourth, this study has limitations inherent to its naturalistic design, including the comparison of pre-pandemic (2019) and pandemic-era (2021) cohorts. The psychological, social, and academic disruptions caused by the pandemic may have influenced the 2021 cohort’s appreciation for online PBL, potentially inflating satisfaction due to the availability of continued education during a crisis rather than reflecting a clear preference for the online format itself.

Additionally, variables such as increased exposure to digital tools, variable home learning conditions, and changes in institutional teaching practices during the pandemic complicate the interpretation of results. Nevertheless, this study’s findings remain relevant and supported by comparable work conducted both during and outside the pandemic. For instance, another study conducted during the pandemic demonstrated enhanced student satisfaction and perceived effectiveness of virtual PBL despite challenges in transitioning[36]. Similarly, a Brazilian study, who examined during the pandemic hybrid formats, reported that well-structured online PBL—with appropriate facilitation—continued to foster student engagement and learning outcomes[37]. Donkin et al. conducted a scoping review of 13 studies on online Case-Based Learning (CBL) in medical education published between 2010 and 2022, including 9 studies during the COVID-19 pandemic and 4 from before[38]. Given the pedagogical similarities between CBL and Problem-Based Learning (PBL)—both relying on collaborative, student-centered case analysis—the findings are highly relevant. The review showed that online formats maintained or improved learning outcomes, despite challenges like communication barriers and internet access. While replication in non-pandemic contexts is needed, our findings contribute to a growing consensus that hybrid or online PBL can address demographic and linguistic barriers in global medical education.

### Future Directions

Future research should focus on fostering collaborations between universities from different countries to implement and study PBL among non-native English-speaking populations. Such initiatives could significantly boost students’ interest and participation by exposing them to diverse educational practices and cultural perspectives. Cross-cultural studies involving diverse student populations are essential to validate our findings and refine PBL strategies to accommodate different linguistic and cultural contexts. Finally, additional efforts should aim at enhancing critical thinking and addressing demographic disparities in PBL satisfaction. As the majority of medical literature is published in English [39], proficiency in this language is essential for future medical doctors. It enables them to critically assess the quality and reliability of their sources, ensuring that their clinical decisions are based on the best available evidence. Moreover, English proficiency allows medical professionals to engage with the latest advancements in the field, participate in international collaborations, and continuously update their knowledge throughout their careers, fostering lifelong learning and evidence-based practice.

## Conclusion

Our findings underscore the potential of online PBL to improve student satisfaction in medical education, particularly for topics requiring high engagement and reflection. By leveraging digital tools and addressing demographic factors, educators can optimize PBL experiences to meet the evolving needs of students. These insights contribute to the growing body of evidence supporting innovative approaches to medical education and highlight the importance of tailoring PBL to diverse learning environments.

## Data Availability

The datasets used and/or analyzed during the current study are available from the corresponding author upon reasonable request.

## Author Contributions

Conceptualization, B.C., B.A. and N.S.; formal analysis, B.C., K.N., T.K., N.T., H.T., H.N., B.A., C.D., I.B.-V., T.Y., N.S. and H.K.; writing—original draft preparation, B.C., K.N., T.K., N.T., H.T., H.N., B.A., C.D., I.B.-V., T.Y., N.S. and H.K.; writing—review and editing, B.C., K.N., T.K., N.T., H.T., H.N., B.A., C.D., I.B.-V., T.Y., N.S. and H.K.; supervision, B.A., N.S. and H.K.; project administration, B.A., N.S. and H.K. All authors have read and agreed to the published version of the manuscript.

## Ethics declarations

All procedures performed in the study involving human participants were in accordance with the ethical standards of the institutional and/or national research committee and with the 1964 Helsinki Declaration and its later amendments or comparable ethical standards. This study was approved by representatives of the ethical committee at Nagoya University (Approval Code: 2021-0482).

## Informed Consent Statement

Details of the study were explained to participants beforehand, and signed consent was obtained from all participants.

